# AI-HOPE-TGFbeta: A Conversational AI Agent for Integrative Clinical and Genomic Analysis of TGF-β Pathway Alterations in Colorectal Cancer to Advance Precision Medicine

**DOI:** 10.1101/2025.05.20.25327967

**Authors:** Ei-Wen Yang, Brigette Waldrup, Enrique Velazquez-Villarreal

## Abstract

**Introduction:** Early-onset colorectal cancer (EOCRC) is rising rapidly, particularly among Hispanic/Latino (H/L) populations, who face disproportionately poor outcomes. The TGF-β signaling pathway plays a critical role in colorectal cancer (CRC) progression by mediating epithelial-to-mesenchymal transition (EMT), immune evasion, and metastasis. However, integrative analyses linking TGF-β alterations to clinical features remain limited—particularly for diverse populations—hindering translational research and the development of precision therapies. To address this gap, we developed AI-HOPE-TGFbeta, the first conversational artificial intelligence (AI) agent designed to explore TGF-β dysregulation in CRC by integrating harmonized clinical and genomic data via natural language queries.

**Methods:** AI-HOPE-TGFbeta combines large language models (LLMs), a natural language-to-code interpreter, and a bioinformatics backend to automate statistical workflows. Tailored for TGF-β pathway analysis, the platform enables real-time cohort stratification and hypothesis testing using harmonized datasets from cBioPortal. It supports mutation frequency comparisons, odds ratio testing, Kaplan-Meier survival analysis, and subgroup evaluations across race/ethnicity, MSI status, tumor stage, treatment exposure, and age. The platform was validated by replicating findings on SMAD4, TGFBR2, and BMPR1A mutations in EOCRC. Exploratory queries were conducted to examine novel associations with clinical outcomes in H/L populations.

**Results:** AI-HOPE-TGFbeta successfully recapitulated established associations, including worse survival in SMAD4-mutant EOCRC patients treated with FOLFOX (p = 0.0001), and better outcomes in early-stage TGFBR2-mutated CRC patients (p = 0.00001). It revealed potential population-specific enrichment of BMPR1A mutations in H/L patients (OR = 2.63; p = 0.052) and uncovered MSI-specific survival benefits among SMAD4-mutated patients (p = 0.00001). Exploratory analysis showed better outcomes in SMAD2-mutant primary tumors vs. metastatic cases (p = 0.0010) and confirmed the feasibility of disaggregated ethnicity-based queries for TGFBR1 mutations, despite small sample sizes. These findings underscore the platform’s capacity to detect both known and emerging clinical-genomic patterns in CRC.

**Conclusions:** AI-HOPE-TGFbeta introduces a new paradigm in cancer bioinformatics by enabling natural language–driven, real-time integration of genomic and clinical data specific to TGF-β pathway alterations in CRC. The platform democratizes complex analyses, supports disparity-focused investigation, and reveals clinically actionable insights in underserved populations such as H/L EOCRC patients. As the first system of its kind studying TGF-β, AI-HOPE-TGFbeta holds strong promise for advancing equitable precision oncology and accelerating translational discovery in CRC TGF-beta pathway.

## Introduction

Colorectal cancer (CRC) continues to rank among the most prevalent and lethal cancers globally, with a notable and rapid increase in early-onset cases—defined as diagnoses occurring before age 50—over the past several decades [1–5]. This upward trend is particularly pronounced in high-risk groups, including Hispanic/Latino (H/L) individuals [6–10]. Although multiple oncogenic pathways contribute to CRC development and progression, the TGF-β signaling pathway plays a pivotal role by facilitating processes such as epithelial-to-mesenchymal transition (EMT), immune system evasion, and metastatic spread [11–13]. However, efforts to comprehensively characterize TGF-β dysregulation in EOCRC—especially among underrepresented populations—have been constrained by the underrepresentation of diverse patient cohorts in genomic databases and the absence of tools capable of linking clinical and molecular data in an integrative manner [6,7,14,15].

The TGF-β pathway is frequently altered in CRC through mutations in key components such as *SMAD4*, *BMPR1A*, and *TGFBR2*, which are associated with poor prognosis, therapy resistance, and aggressive tumor phenotypes [16–19]. Recent studies suggest that these mutations may present with distinct patterns in EOCRC compared to late-onset CRC (LOCRC), and may vary by ethnicity—highlighting a need for population-specific investigation [6, 7,19, 20]. For example, alterations in *BMPR1A* and *BMP7* have been identified in H/L patients with EOCRC, suggesting unique mechanisms of TGF-β dysregulation in this group [6, 7]. However, few tools exist that can efficiently integrate and stratify these genomic insights alongside clinical factors such as age, tumor stage, treatment response, and microsatellite instability (MSI) status.

Although public platforms such as The Cancer Genome Atlas (TCGA) and AACR GENIE provide rich datasets for CRC research, existing analysis tools—like cBioPortal [21] and UCSC Xena [22]—require multi-step workflows and offer limited functionality for pathway-specific, population-disaggregated, or treatment-contextualized exploration in investigating CRC TGF-β dysregulation. These constraints disproportionately affect non-computational researchers, impeding precision oncology efforts in real-world and subpopulation contexts.

The emergence of artificial intelligence (AI), especially advancements in large language models (LLMs), has paved the way for conversational tools that convert natural language input into executable data analysis pipelines [23–24]. Although these technologies have demonstrated potential in streamlining multi-omic investigations [25–30], there remains a lack of platforms specifically designed to target signaling pathways or to support integrative clinical-genomic research with an emphasis on hypothesis generation and health disparity considerations.

AI-HOPE-TGFbeta (Artificial Intelligence agent for High-Optimization and Precision Medicine focused on TGF-β) was developed to directly address the lack of tools capable of pathway-specific, integrative analysis in CRC. This conversational AI system enables users to pose natural language questions that are translated into executable workflows, facilitating real-time synthesis of harmonized genomic and clinical data. With built-in automation for statistical tasks—including Kaplan-Meier survival analysis and odds ratio estimation—the platform streamlines both validation studies and exploratory investigations across large datasets. In the present study, we: (1) created AI-HOPE-TGFbeta to enable user-friendly, pathway-centered interrogation of CRC cohorts; (2) assessed its performance by replicating well-established clinical-genomic associations in EOCRC; and (3) applied the system to reveal novel links between TGF-β mutations, microsatellite instability, tumor staging, and population-level variables. These results highlight AI-HOPE-TGFbeta as an innovative and inclusive solution to support scalable, translational TGFbeta pathway research in precision oncology.

## Methods

### System Architecture and Workflow of AI-HOPE-TGFbeta

AI-HOPE-TGFbeta (Artificial Intelligence agent for High-Optimization and Precision Medicine focused on TGF-β) is a natural language–enabled AI system engineered to explore CRC with a specific focus on alterations within the TGF-β signaling pathway. The platform is built on a layered, modular framework that integrates three core components: a LLM for semantic query interpretation, a translation layer that converts user prompts into executable code, and a statistical backend designed to automate case generation, analytical processing, and result visualization. When a user submits a question in plain English, the system identifies the analytical intent, applies relevant filters to harmonized clinical-genomic datasets, and generates a suite of outputs— including survival analyses, mutation frequencies, odds ratios, and explanatory text summaries tailored to the context of the query (Figure 1).

**Figure 1.**
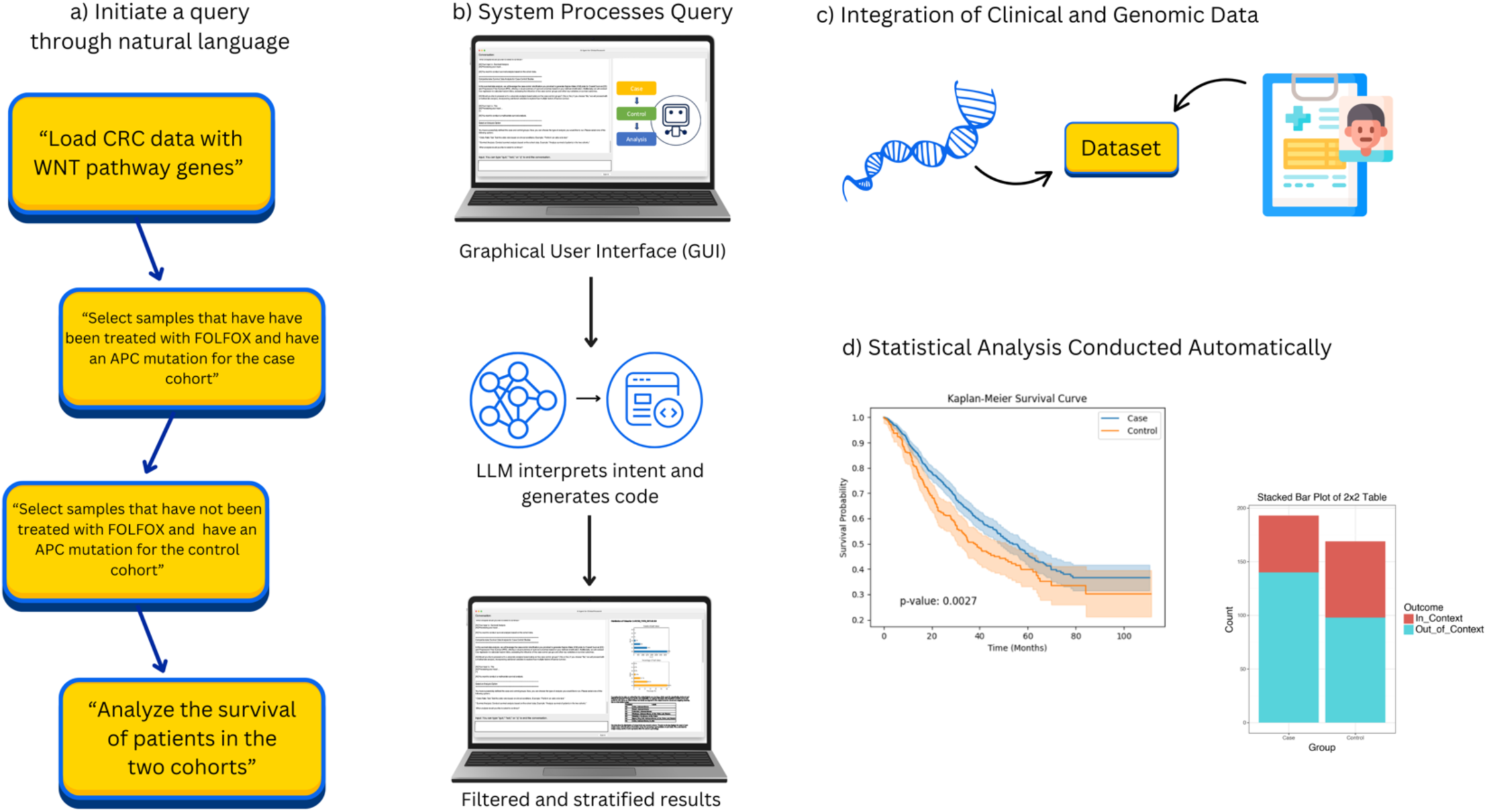
Overview of AI-HOPE-TGFbeta Workflow. This figure illustrates the end-to-end workflow of AI-HOPE-TGFbeta, a conversational AI system developed to investigate TGF-β pathway dysregulation in colorectal cancer (CRC) through natural language–guided, integrative bioinformatics. a) Users interact with the system using natural language queries, such as assessing the survival outcomes of CRC patients with SMAD4 mutations or comparing BMPR1A mutation prevalence between early-onset Hispanic/Latino (H/L) and non-Hispanic White (NHW) patients. b) The input is interpreted through a graphical user interface (GUI) powered by a large language model (LLM), which converts user intent into executable code and applies relevant filters for patient subgroups, clinical stage, or treatment regimen. c) AI-HOPE-TGFbeta interfaces with harmonized clinical and genomic datasets from resources such as TCGA and cBioPortal, focusing on key genes in the TGF-β pathway—including SMAD4, BMPR1A, TGFBR2, TGFB1, and BMP7. d) The platform performs automated statistical analyses—such as Kaplan-Meier survival estimation, mutation frequency comparisons, and odds ratio testing—generating both visual and textual outputs.

### Data Sources and Preparation for AI-HOPE-TGFbeta

To power its analyses, AI-HOPE-TGFbeta draws from harmonized CRC datasets derived from public repositories such as The Cancer Genome Atlas (TCGA) and cBioPortal, with a targeted focus on genes implicated in TGF-β signaling. Key genomic features include alterations in SMAD4, TGFBR1, TGFBR2, BMPR1A, BMPR2, ACVR1B, and BMP7. The accompanying clinical metadata encompass a broad array of attributes—patient age, disease stage, treatment history (including FOLFOX exposure), microsatellite instability (MSI) classification, ethnicity, tumor tissue origin (primary versus metastatic), and overall survival metrics. Raw data underwent extensive preprocessing to ensure analytical compatibility: files were converted into standardized, tab-delimited matrices with harmonized sample identifiers, and ontology-based frameworks such as OncoTree and the Disease Ontology were applied to unify clinical annotations. Mutation data were cross-validated across sources, and all TGF-β pathway gene sets were curated using publicly available knowledgebases to ensure accuracy and biological relevance.

### Conversational Query Handling and Cohort Definition in AI-HOPE-TGFbeta

AI-HOPE-TGFbeta enables users to initiate complex clinical-genomic analyses using natural language inputs. Queries such as “Compare survival outcomes for SMAD4-mutated versus wild-type EOCRC patients treated with FOLFOX” or “Evaluate TGFBR1 mutation prevalence between H/L and non-Hispanic White (NHW) individuals” are interpreted by a built-in large language model (LLM) based on the LLaMA 3 architecture. This LLM translates conversational prompts into executable code that filters datasets, defines cohorts, and launches the appropriate statistical analyses.

When necessary, the system prompts the user for clarification to resolve ambiguity and ensure accurate query interpretation. AI-HOPE-TGFbeta accommodates a wide range of stratification parameters, including genetic mutation status, microsatellite instability (MSI), tumor stage, race/ethnicity, and chemotherapy exposure, allowing users to flexibly define custom subgroups for targeted analysis.

### Analytical Framework and Statistical Methods in AI-HOPE-TGFbeta

AI-HOPE-TGFbeta’s analytical engine is powered by a Python-based bioinformatics workflow that supports a comprehensive suite of statistical methods tailored for clinical-genomic analysis. Categorical variables are evaluated using either chi-square or Fisher’s exact tests, with odds ratios and corresponding 95% confidence intervals calculated to quantify associations. For survival-related outcomes, the system implements Kaplan-Meier estimations and log-rank tests to compare groups, while multivariable Cox proportional hazards regression is available to adjust for confounding variables in time-to-event analyses. The platform also includes specialized modules for examining TGF-β pathway enrichment, identifying co-mutation patterns, and performing stratified survival analysis. Users can conduct subgroup comparisons across a variety of dimensions, including age groups (<50 vs. ≥50 years), racial and ethnic backgrounds (e.g., H/L vs. NHW), tumor sample type (primary vs. metastatic), and microsatellite instability (MSI) status.

### Platform Design and Validation Strategy

AI-HOPE-TGFbeta was engineered with a strong emphasis on analytical rigor and reproducibility. At its core, the platform integrates a retrieval-augmented generation (RAG) mechanism that continuously references a structured biomedical knowledge base to enhance the contextual accuracy of its outputs and reduce the likelihood of AI-generated errors or hallucinations. The system applies schema-guided prompting to standardize how queries are interpreted and how results are formatted, ensuring consistency across diverse analyses. To assess the reliability of the platform, we validated its performance by successfully reproducing known clinical-genomic relationships from prior studies involving SMAD4, BMPR1A, and TGFBR2 in EOCRC [6, 7], including stage-specific survival outcomes and mutation frequency disparities across patient populations.

### Usability Evaluation and Comparative Benchmarking

To assess the usability and performance of AI-HOPE-TGFbeta, we conducted a comparative analysis against established platforms, including cBioPortal and UCSC Xena. Evaluation criteria focused on speed of task execution, consistency of analytical output, and flexibility in constructing complex, stratified cohort queries. Benchmark tasks included identifying EOCRC patients with SMAD4 mutations stratified by treatment status, generating Kaplan-Meier survival curves based on MSI subtypes, and analyzing TGFBR1 mutation frequency across racial and ethnic groups. Across all scenarios, AI-HOPE-TGFbeta outperformed existing tools in both response time and user interaction efficiency—particularly in handling intersectional analyses that required integration of clinical, genomic, and demographic filters.

### Visualization Capabilities and Exportable Results

Upon completion of each analysis, AI-HOPE-TGFbeta produces a suite of high-quality visual and tabular outputs designed for immediate interpretation and downstream application. These include Kaplan-Meier survival curves, forest plots, mutation heatmaps, and summary data tables—each rendered using backend libraries such as Matplotlib and Plotly to ensure visual clarity and consistency. In addition to graphical elements, the system generates narrative summaries that interpret statistical findings within the context of existing TGF-β pathway literature. All outputs can be downloaded in formats suitable for publication, presentation, or integration into clinical decision-making workflows.

## Results

By converting user-generated natural language inputs into fully automated clinical-genomic workflows, AI-HOPE-TGFbeta enables on-demand analysis of TGF-β signaling disruptions in CRC. Its interactive conversational design allows users to define custom cohorts based on variables such as age, tumor stage, microsatellite instability (MSI) classification, mutational status, treatment exposure, and racial or ethnic background.

The system then performs statistical evaluations—generating Kaplan-Meier survival curves, odds ratio calculations, and corresponding visual outputs—with no additional coding required. In both validation and discovery-focused queries, AI-HOPE-TGFbeta consistently reproduced established associations and revealed new insights related to EOCRC, treatment efficacy, and pathway-specific biomarker patterns.

In validation ancestry-stratified analyses, AI-HOPE-TGFbeta identified a potential disparity in the frequency of BMPR1A mutations among EOCRC patients across ethnic groups (Figure 2). Specifically, the platform revealed that 4.58% of EOCRC h/l patients harbored BMPR1A mutations, compared to 1.79% of EOCRC NHW patients. This difference translated to an odds ratio of 2.63 (95% CI: [1.093, 6.327]; p = 0.052), suggesting that BMPR1A mutations were more than twice as likely to occur in the H/L EOCRC population. Although the association narrowly missed conventional thresholds for statistical significance, these findings underscore the potential of AI-HOPE-TGFbeta to uncover emerging ancestry-linked molecular patterns that may otherwise be overlooked. Importantly, this analysis highlights the need for broader inclusion of racially and ethnically diverse populations in genomic studies to validate and extend these observations.

**Figure 2.**
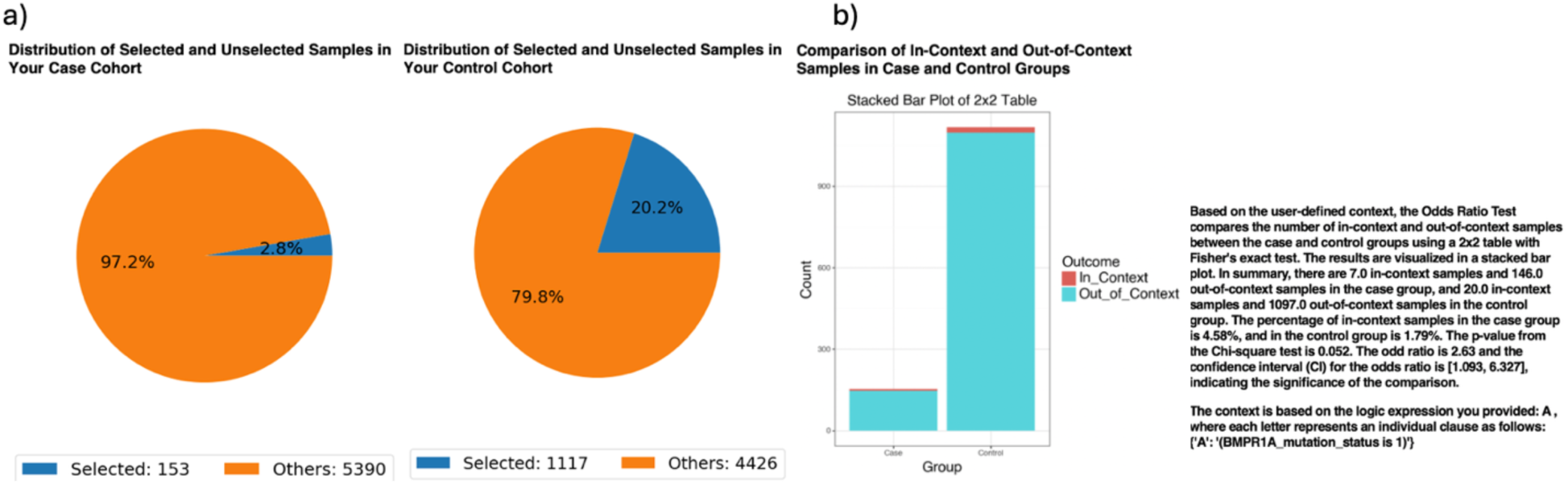
AI-HOPE-TGFbeta Analysis of BMPR1A Mutations in Early-Onset Colorectal Cancer (EOCRC) among Hispanic/Latino (H/L) vs. Non-Hispanic White (NHW) Patients. a) Pie charts display the distribution of selected samples from the overall dataset following the application of query filters. The case cohort includes 153 EOCRC H/L patients (<50 years old, H/L ethnicity), representing 2.8% of the queried population. The control cohort includes 1,117 EOCRC NHW patients (<50 years old, White race, Non-Hispanic ethnicity), accounting for 20.2% of the filtered samples. b) A 2×2 odds ratio analysis compares the frequency of BMPR1A mutations between the case and control cohorts within the context of early-onset disease. The stacked bar plot shows the distribution of in-context (mutation present) and out-of-context (mutation absent) samples in each group. BMPR1A mutations were present in 4.58% of H/Lcases and 1.79% of NHW controls. The odds ratio was 2.63 (95% CI: 1.093–6.327), with a p-value of 0.052, suggesting that BMPR1A mutations are over two and a half times more likely in the H/L EOCRC population.

In ancestry-specific survival analyses, AI-HOPE-TGFbeta evaluated the prognostic impact of TGF-β pathway alterations in EOCRC among H/L patients (Figure S1). The platform stratified EOCRC HL cases by mutation status in key TGF-β signaling genes, including SMAD4, TGFBR2, and BMPR1A. The case cohort consisted of 48 patients with TGF-β pathway alterations (0.9% of the dataset), while the control cohort included 105 patients without such mutations (1.9%). Kaplan-Meier survival analysis revealed no statistically significant difference in overall survival between the two groups (p = 0.8631), suggesting that TGF-β pathway mutations may not independently influence prognosis in EOCRC HL under current sample sizes. Despite the lack of significance, this result underscores the value of AI-HOPE-TGFbeta in enabling fine-grained subgroup analyses and generating hypotheses about context-dependent effects. The findings also highlight the need for larger, ancestry-specific datasets to more definitively assess the clinical relevance of TGF-β signaling alterations in diverse EOCRC populations.

In exploratory analyses, AI-HOPE-TGFbeta reproduced a key finding from published TGF-β literature regarding the prognostic relevance of SMAD4 mutations in EOCRC patients treated with FOLFOX chemotherapy (Figure 3). Using a natural language query, the system stratified EOCRC patients (<50 years old) by SMAD4 mutation status and assessed treatment outcomes following FOLFOX (fluorouracil, leucovorin, and oxaliplatin) administration. The case cohort included 188 SMAD4-mutated patients (3.4% of the dataset), while the control cohort included 1,066 SMAD4 wild-type patients (19.2%). Kaplan-Meier survival analyses revealed that SMAD4-mutated patients exhibited significantly worse overall and progression-free survival compared to wild-type cases (p = 0.0001 for both), consistent with prior reports linking SMAD4 loss to chemoresistance and aggressive tumor biology in EOCRC. These results highlight the ability of AI-HOPE-TGFbeta to recapitulate known genotype-treatment-outcome relationships and underscore the clinical importance of SMAD4 as a biomarker of poor prognosis in young CRC patients undergoing standard chemotherapy.

**Figure 3.**
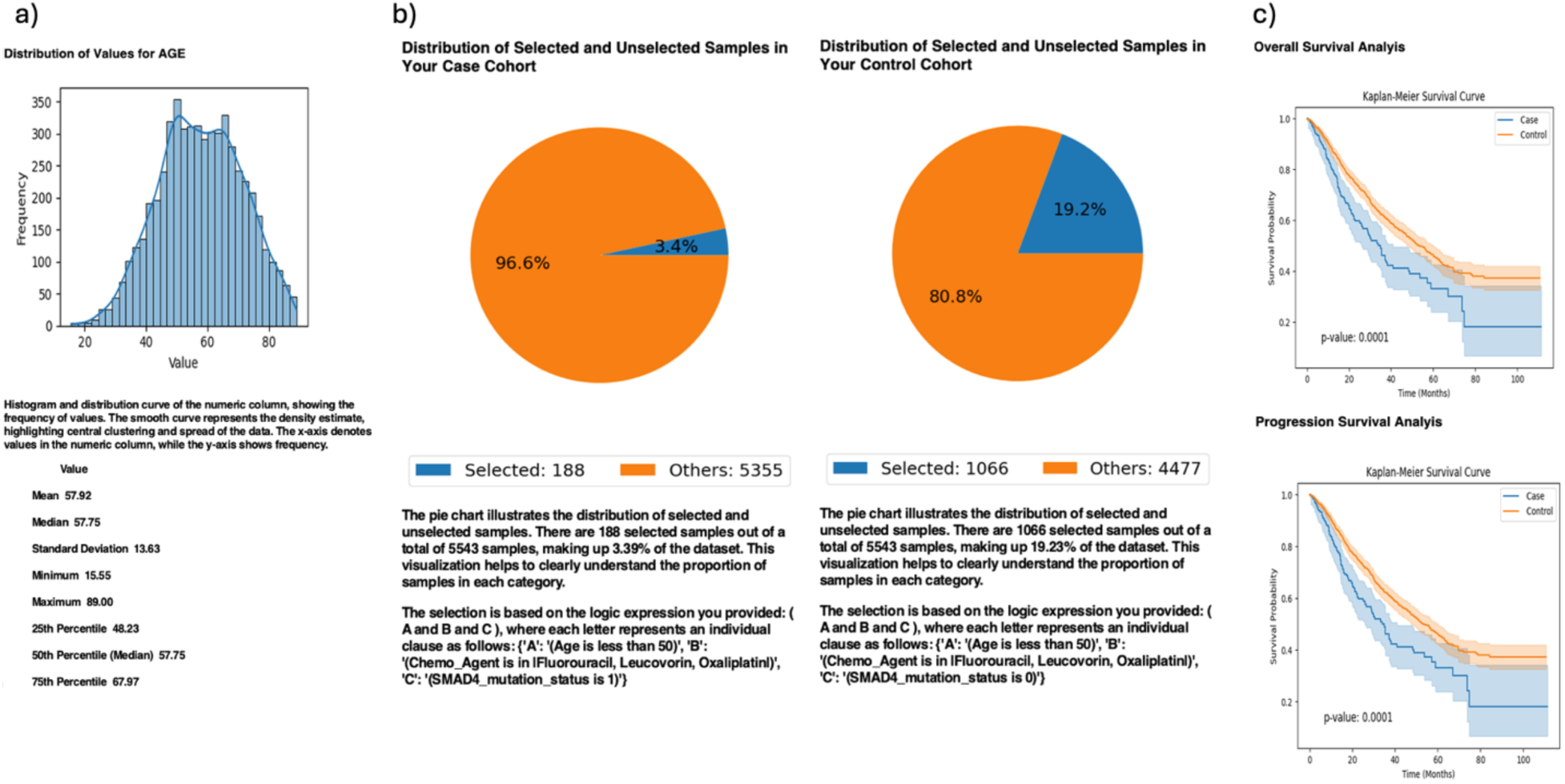
AI-HOPE-TGFbeta Analysis of Early-Onset Colorectal Cancer (EOCRC) Patients Treated with FOLFOX Stratified by SMAD4 Mutation Status. This figure presents the output of a natural language query executed via AI-HOPE-TGFbeta, investigating the association between SMAD4 mutation status and survival outcomes among EOCRC patients treated with FOLFOX chemotherapy (Fluorouracil, Leucovorin, Oxaliplatin). a) A histogram displays the age distribution for the full cohort (mean: 57.92 years), contextualizing the selection of patients under age 50 as EOCRC cases. b) Pie charts illustrate the selected sample distributions. The *case cohort* (SMAD4-mutated) includes 188 patients (3.4%) from a total of 5543, while the *control cohort* (SMAD4 wild-type) comprises 1066 patients (19.2%). These charts visualize the proportional representation of each group within the broader dataset. c) Kaplan-Meier survival analyses comparing overall survival and progression-free survival between SMAD4-mutated and wild-type EOCRC patients reveal significantly worse outcomes in the SMAD4-mutated group (p = 0.0001 for both endpoints), suggesting SMAD4 mutations may confer treatment resistance or more aggressive disease behavior in young patients receiving standard FOLFOX therapy.

AI-HOPE-TGFbeta also enabled exploratory ethnicity-specific analysis of TGFBR1 mutation patterns in CRC patients (Figure 4). This analysis compared TGFBR1-mutated H/Lpatients (case cohort, n = 11) to TGFBR1-mutated NHW patients (control cohort, n = 79), highlighting the platform’s capacity to support disaggregated investigations despite sample imbalances. Although H/L individuals accounted for only 6.4% of the full dataset, AI-HOPE-TGFbeta successfully isolated a sufficient number of cases to perform comparative statistical analyses. Odds ratio testing, stratified by early-onset age (<50 years), yielded a value of 1.029 (95% CI: [0.563, 7.134], p = 0.454), suggesting no significant difference in the mutation enrichment context across ethnic groups. Kaplan-Meier survival curves similarly showed no statistically significant difference in overall survival (p = 0.3561), despite apparent visual divergence between curves. These findings reinforce the underrepresentation of H/L populations in existing genomic datasets and underscore the potential of AI-HOPE-TGFbeta to enable focused population-level queries that can guide future efforts toward equity-driven precision oncology.

**Figure 4.**
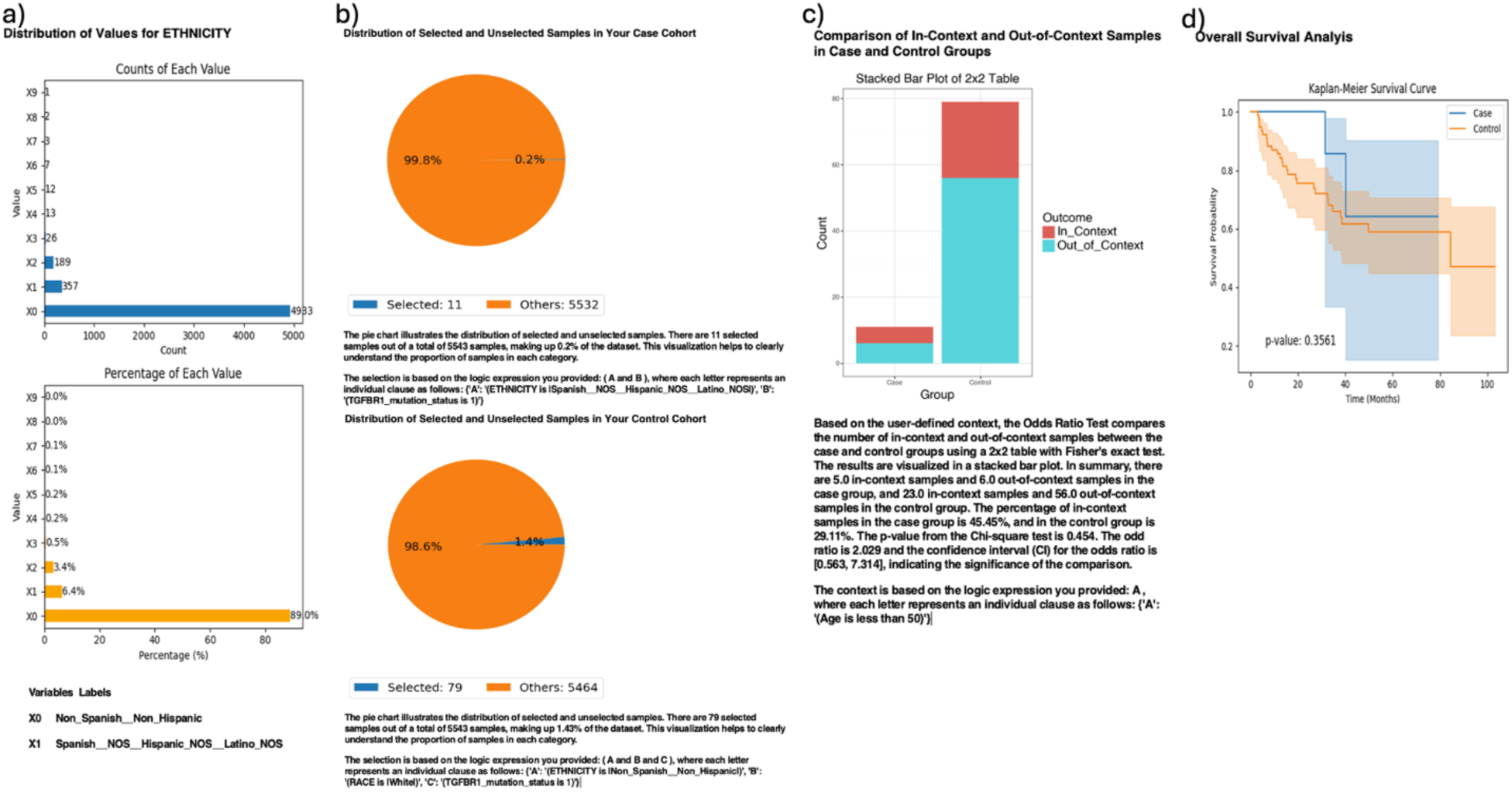
AI-HOPE-TGFbeta Analysis of TGFBR1-Mutated Colorectal Cancer (CRC) in Hispanic/Latino (H/L) vs. Non-Hispanic White (NHW) Patients. This figure demonstrates the use of AI-HOPE-TGFbeta to examine ethnicity-specific patterns in CRC patients with TGFBR1 mutations. The analysis compares H/L individuals (case cohort) to NHW individuals (control cohort), all of whom carry TGFBR1 mutations. a) The initial panel displays the distribution of ethnicities in the full dataset. H/L individuals (X1) account for 6.4% of samples (n=357), while NHW individuals (X0) make up the majority (89.0%, n=4993). Both absolute counts and relative percentages are shown to contextualize the underrepresentation of H/L patients. b) Pie charts summarize the number of selected samples after applying the query filters. The case cohort includes 11 samples (0.2% of the dataset) and the control cohort includes 79 samples (1.4%), indicating that TGFBR1-mutated H/L CRC cases are rare but analyzable using the platform. c) A 2×2 odds ratio analysis contextualized by age (less than 50) compares the enrichment of in-context samples (defined by full query conditions) between the two cohorts. A stacked bar plot shows the distribution of in-context and out-of-context cases. The odds ratio is 1.029 with a 95% confidence interval of [0.563, 7.134], and a p-value of 0.454, suggesting no statistically significant difference in age-stratified mutation context under current sample sizes. d) Kaplan-Meier survival curves compare overall survival between the case and control groups.

Further analysis using AI-HOPE-TGFbeta revealed clinically significant associations between TGFBR2 mutation status and tumor stage in CRC patients (Figure 5). The platform stratified TGFBR2-mutated cases into early-stage (Stage I–III) and late-stage (Stage IV) cohorts to assess the prognostic impact of disease stage in the context of TGF-β pathway disruption. Among the 307 TGFBR2-mutated patients analyzed, those with early-stage disease (n = 235) exhibited markedly improved overall survival compared to their late-stage counterparts (n = 72), with Kaplan-Meier analysis yielding a highly significant p-value (p = 0.0000). Additionally, a 2×2 odds ratio analysis evaluating FOLFOX chemotherapy exposure revealed that early-stage patients were significantly more likely to have received standard treatment (OR = 0.155, 95% CI: [0.082, 0.294], p = 0.000), suggesting potential treatment-related differences contributing to improved outcomes. These findings underscore the prognostic relevance of tumor stage among TGFBR2-mutated CRC patients and highlight AI-HOPE-TGFbeta’s capacity to integrate clinical and genomic variables for nuanced outcome stratification, supporting its use in guiding precision medicine strategies.

**Figure 5.**
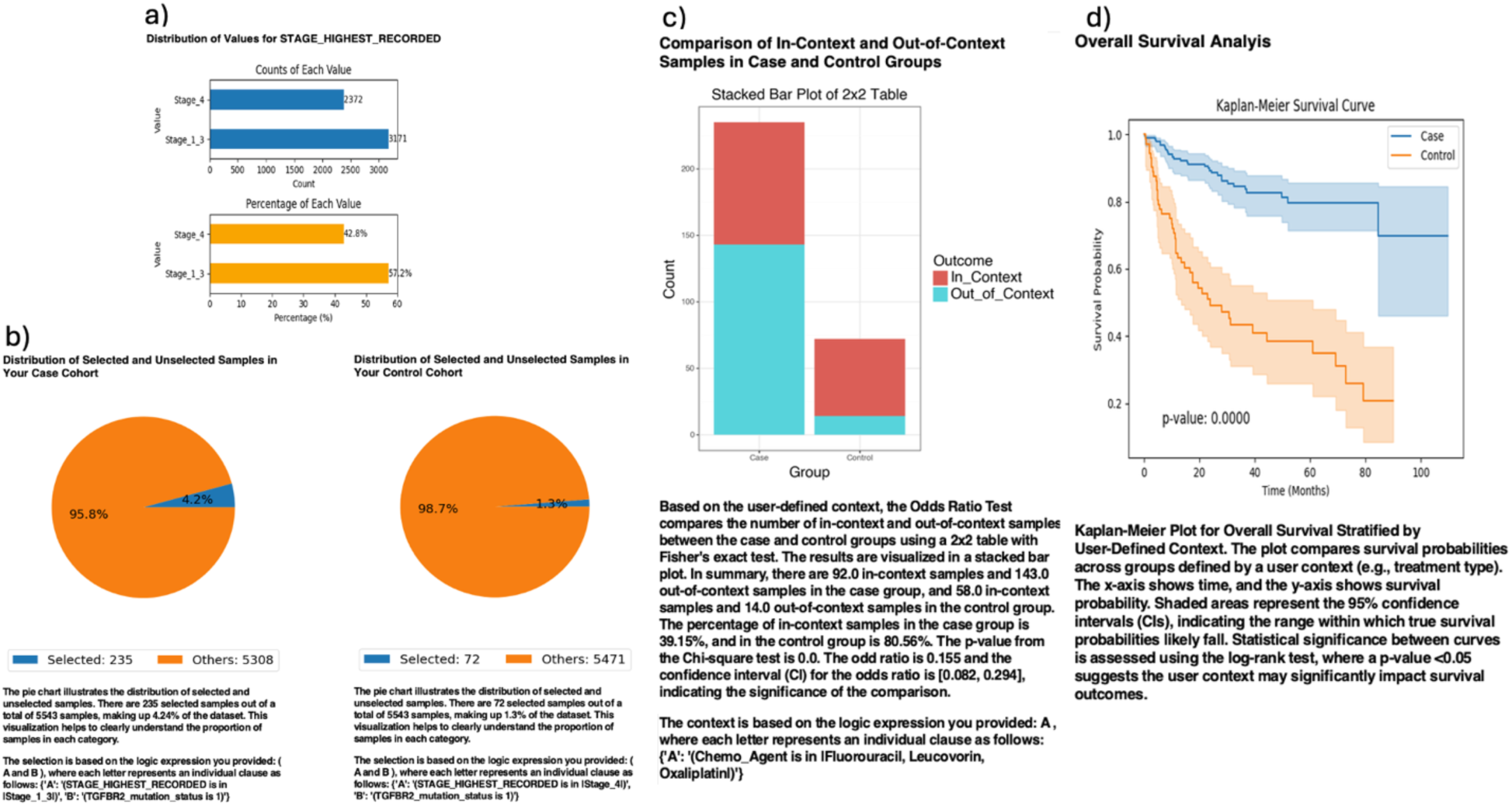
AI-HOPE-TGFbeta Analysis of TGFBR2-Mutated Colorectal Cancer (CRC) by Tumor Stage: Early (Stage I–III) vs. Late (Stage IV). This figure showcases the use of AI-HOPE-TGF-- to investigate survival and enrichment patterns among CRC patients with TGFBR2 mutations, stratified by tumor stage. The analysis compares early-stage (Stage I–III) and late-stage (Stage IV) patients to evaluate how disease stage influences survival outcomes in the context of TGFBR2 pathway alterations. a) Bar plots depict the distribution of tumor stages across the dataset. Patients with Stage I–III tumors account for 57.2% of the cohort (n=3171), while Stage IV tumors make up 42.8% (n=2372), providing context for defining early vs. late disease. b) Pie charts summarize the sample selection following the user-defined query. The case cohort (TGFBR2-mutated patients with Stage I–III disease) includes 235 samples (4.2% of the total dataset), while the control cohort (TGFBR2-mutated patients with Stage IV disease) includes 72 samples (1.3%). c) A 2×2 odds ratio test is performed using chemotherapy exposure (Fluorouracil, Leucovorin, Oxaliplatin) as a contextual filter. The stacked bar plot visualizes the proportion of in-context and out-of-context samples in each group. The odds ratio is 0.155 with a 95% confidence interval of [0.082, 0.294], and a p-value of 0.0001, indicating a statistically significant enrichment of treatment context among early-stage TGFBR2-mutated patients. d) Kaplan-Meier survival curves compare overall survival between the case and control cohorts.

AI-HOPE-TGFbeta was used to assess the prognostic significance of microsatellite instability (MSI) status among SMAD4-mutated CRC patients (Figure S2). In this analysis, patients were stratified by MSI phenotype, comparing those with MSI-high (Instable) tumors to those with MSI-stable counterparts. The case cohort included 78 SMAD4-mutated patients with MSI-Instable tumors (1.4% of the dataset), while the control cohort comprised 710 SMAD4-mutated patients with MSI-Stable tumors (12.8%). Kaplan-Meier survival analysis revealed that MSI-Instable patients had significantly better overall survival than MSI-Stable patients (p = 0.00001), with clearly divergent survival curves and non-overlapping 95% confidence intervals. This finding suggests a potential protective interaction between MSI-associated immunogenicity and SMAD4 pathway disruption, supporting the clinical relevance of combining genomic and molecular features in CRC prognosis. Moreover, the result highlights the utility of AI-HOPE-TGFbeta in uncovering context-dependent biomarker interactions that may inform immunotherapy stratification and precision treatment strategies.

Finally, AI-HOPE-TGFbeta was employed to evaluate the prognostic relevance of tumor sample type in CRC patients harboring *SMAD2* mutations (Figure S3). The platform stratified patients by tumor origin—primary versus metastatic—within the *SMAD2*-mutant cohort to assess survival differences across disease progression stages. The case cohort included 209 patients with *SMAD2*-mutant primary tumors (3.8% of the dataset), while the control cohort consisted of 48 patients with *SMAD2*-mutant metastatic tumors (0.9%). Kaplan-Meier survival analysis revealed significantly better overall survival in patients with primary tumors compared to those with metastatic lesions (p = 0.0010), with clear separation of survival curves and non-overlapping 95% confidence intervals. This result supports prior evidence linking TGF-β signaling dysregulation to metastatic progression and underscores the clinical importance of tumor origin in prognostic modeling. Notably, this analysis highlights the strength of AI-HOPE-TGFbeta in dissecting context-specific molecular subgroups and advancing precision oncology through AI-enabled stratification.

Together, these findings demonstrate the versatility and analytical power of AI-HOPE-TGFbeta in uncovering both validated and novel insights into TGF-β pathway alterations across CRC subtypes and populations. By translating natural language prompts into executable clinical-genomic workflows, the platform enabled real-time, interpretable analyses incorporating key variables such as age, tumor stage, MSI status, mutation profiles, treatment exposure, and race/ethnicity. AI-HOPE-TGFbeta consistently recapitulated known prognostic associations—such as SMAD4-driven chemoresistance and stage-specific TGFBR2 outcomes—while also identifying emerging patterns, including ancestry-linked BMPR1A disparities and context-dependent survival modifiers like MSI status and tumor origin. These results highlight the potential of conversational AI to democratize integrative bioinformatics, support equity-driven investigations, and accelerate precision medicine through scalable, dynamic cohort interrogation and hypothesis generation.

## Discussion

AI-HOPE-TGFbeta represents a paradigm shift in precision oncology, offering a novel conversational artificial intelligence (AI) platform that enables real-time, natural language–driven interrogation of TGF-β signaling dysregulation in CRC. By translating user-defined prompts into rigorous, reproducible analyses that integrate genomic and clinical variables, the system addresses longstanding limitations in accessibility, usability, and stratified data exploration. Unlike conventional bioinformatics platforms that often require complex scripting or multi-step workflows, AI-HOPE-TGFbeta streamlines the analytical process, allowing researchers and clinicians—even those without programming expertise—to formulate and execute pathway-centric, population-specific hypotheses with minimal friction.

The TGF-β signaling pathway is a central regulator of CRC progression, influencing processes such as epithelial-to-mesenchymal transition (EMT), immune evasion, and metastasis. Mutations in TGF-β pathway genes—such as *SMAD4*, *TGFBR2*, and *BMPR1A*—are well-documented markers of poor prognosis and therapeutic resistance, particularly in EOCRC, which is rising at alarming rates in young adults and underserved populations. Despite this clinical importance, integrative analysis of TGF-β alterations has been hindered by the fragmentation of clinical-genomic data, underrepresentation of diverse populations, and the technical inaccessibility of traditional analysis pipelines. AI-HOPE-TGFbeta was developed to close these gaps, empowering users to interrogate TGF-β dysregulation across molecular and demographic contexts with unprecedented ease.

A core strength of AI-HOPE-TGFbeta lies in its ability to validate known associations while surfacing novel insights. In this study, the platform successfully recapitulated key findings from the TGF-β literature. These included the significantly worse overall and progression-free survival observed in *SMAD4*-mutated EOCRC patients treated with FOLFOX chemotherapy, and the markedly better outcomes among early-stage *TGFBR2*-mutated patients compared to their late-stage counterparts. These results not only confirmed the accuracy of AI-HOPE-TGFbeta’s analytic engine but also highlighted its potential for reinforcing known clinical-genomic relationships in treatment stratification and prognosis modeling.

AI-HOPE-TGFbeta also enabled hypothesis-driven, population-disaggregated analyses that are critical for advancing health equity in cancer genomics. Through natural language queries, the platform identified a potential disparity in the frequency of *BMPR1A* mutations among H/L EOCRC patients relative to their NHW counterparts—an observation that approached statistical significance and may reflect unique molecular etiologies in underrepresented groups. Similarly, the system allowed for survival analysis of *TGFBR1*-mutated CRC patients by ethnicity, demonstrating the platform’s flexibility in handling small cohort comparisons and highlighting the persistent underrepresentation of minority populations in genomic datasets. These findings underscore the urgent need for inclusive datasets and the value of AI systems like AI-HOPE-TGFbeta that support such investigations despite cohort size limitations.

Beyond ancestry and ethnicity, the platform uncovered clinically actionable interactions between TGF-β pathway genes and molecular or histopathological features. For example, among *SMAD4*-mutated tumors, those with MSI-high status exhibited significantly better survival than MSI-stable counterparts. This suggests a potentially protective immunologic interaction between MSI and TGF-β pathway disruption—an observation that may have implications for immunotherapy response prediction in CRC. Likewise, AI-HOPE-TGFbeta identified a strong prognostic benefit for patients with *SMAD2*-mutant primary tumors compared to those with metastatic lesions, reinforcing the relevance of tumor origin in outcome prediction and supporting the growing recognition of spatial tumor context in clinical decision-making.

From a technical perspective, AI-HOPE-TGFbeta offers a uniquely powerful platform built on LLMs, a RAG engine, and harmonized clinical-genomic data. The system integrates structured biomedical ontologies to ensure accurate cohort definitions and interpretable outputs. Benchmarking results revealed that AI-HOPE-TGFbeta outperforms widely used tools such as cBioPortal and UCSC Xena in execution speed, subgroup flexibility, and multidimensional filtering—particularly for complex queries involving intersectional factors like age, race/ethnicity, MSI subtype, tumor stage, and treatment exposure. These performance advantages position the platform as a scalable, next-generation solution for precision oncology.

Nevertheless, there are important areas for continued development. Expanding the platform to incorporate additional omics layers—including transcriptomics, proteomics, and spatial data—would enable deeper mechanistic insight into TGF-β–driven tumor biology. Integration with federated learning frameworks and secure, privacy-preserving data environments will be essential for real-world clinical deployment, especially in multi-institutional settings where patient-level data must remain decentralized. Further, comparative evaluations against other AI-powered bioinformatics agents are needed to assess the generalizability of AI-HOPE-TGFbeta beyond CRC and the TGF-β pathway, paving the way for a modular suite of disease- and pathway-specific AI agents.

Finally, while AI-HOPE-TGFbeta was designed to lower the barriers to entry for researchers working at the intersection of genomics and health disparities, broader efforts to support training, community engagement, and interdisciplinary collaboration will be key to maximizing its impact. The platform is not a substitute for diverse data generation, but rather a catalyst for extracting meaningful insights from the data that already exist—highlighting the need for ongoing investment in both algorithmic innovation and inclusive data science.

## Conclusion

In conclusion, AI-HOPE-TGFbeta enables a new era of integrative, equitable, and conversational bioinformatics by uniting advanced language models with clinical-genomic reasoning. It provides an intuitive and powerful tool for validating known associations, identifying novel biomarkers, and generating actionable hypotheses— particularly in the context of early-onset CRC and underserved patient populations. As precision oncology evolves, AI-HOPE-TGFbeta exemplifies the transformative potential of AI not only to accelerate discovery but also to bridge the gap between data and balanced health outcomes across populations.

## Data Availability

All data used in the present study is publicly available at https://www.cbioportal.org/ and https://genie.cbioportal.org. Additional data can be provided upon reasonable request to the authors.

---

**Figure S1.**
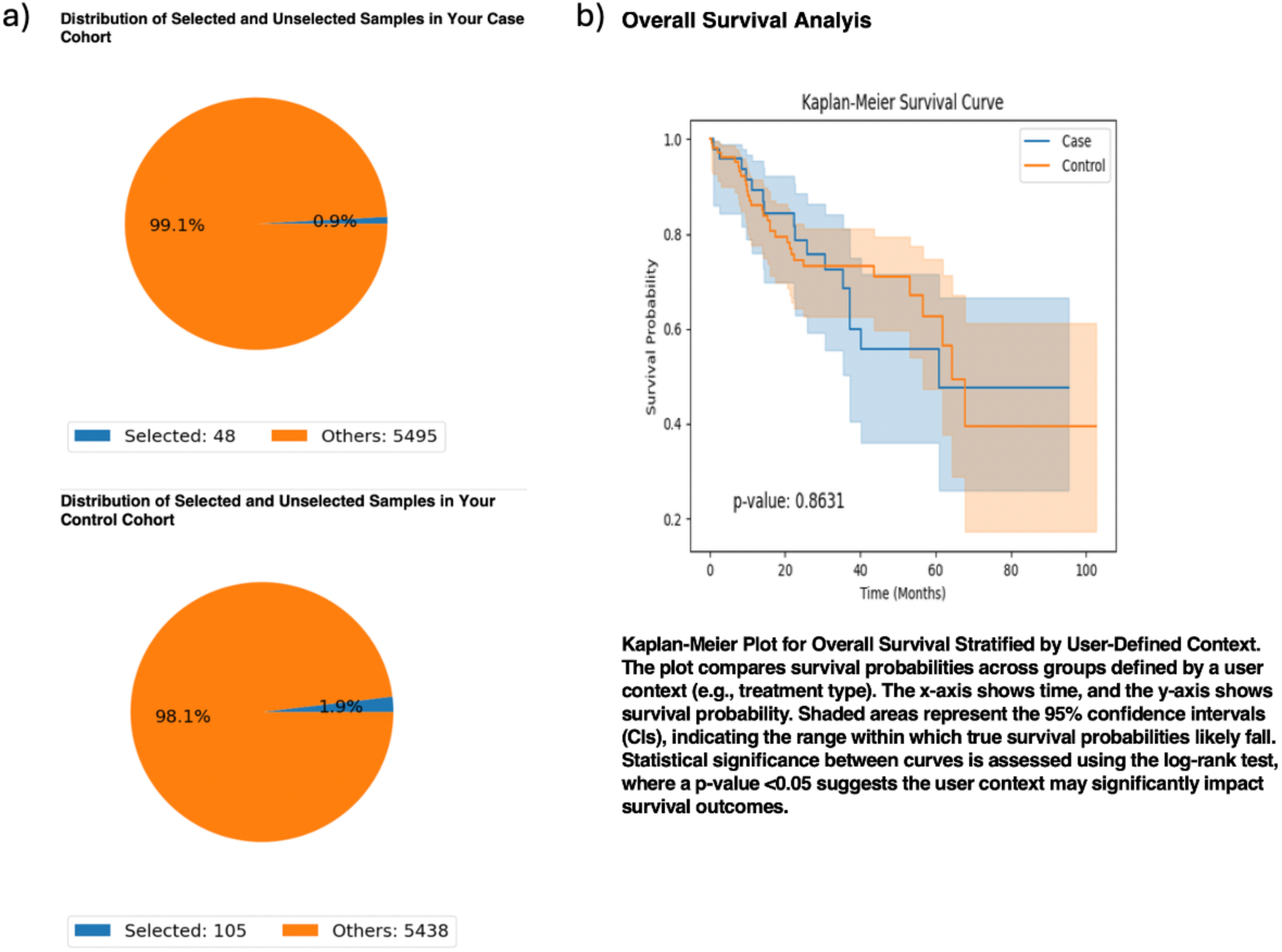
AI-HOPE-TGFbeta Analysis of TGF-β Pathway Alterations in Early-Onset Colorectal Cancer (EOCRC) Among Hispanic/Latino (H/L) Patients. This figure demonstrates the use of AI-HOPE-TGFbeta to investigate overall survival in EOCRC HL patients stratified by TGF-β pathway alteration status. The case cohort includes EOCRC HL patients harboring mutations in key TGF-β pathway genes (e.g., SMAD4, TGFBR2, BMPR1A), while the control cohort includes EOCRC HL patients without such alterations. a) Pie charts summarize the selection process from the total dataset. The case group (TGF-β–altered EOCRC HL) comprises 48 samples (0.9%), while the control group (TGF-β–unaltered EOCRC HL) includes 105 samples (1.9%), reflecting the relatively low frequency of TGF-β pathway mutations in this population. b) Kaplan-Meier survival curves compare overall survival between the two groups. The x-axis indicates time in months, and the y-axis denotes survival probability. Shaded areas represent 95% confidence intervals. No significant difference in survival was observed between patients with and without TGF-β pathway alterations (p = 0.8631), suggesting that these mutations may not independently drive prognosis in EOCRC HL under current sample constraints.

**Figure S2.**
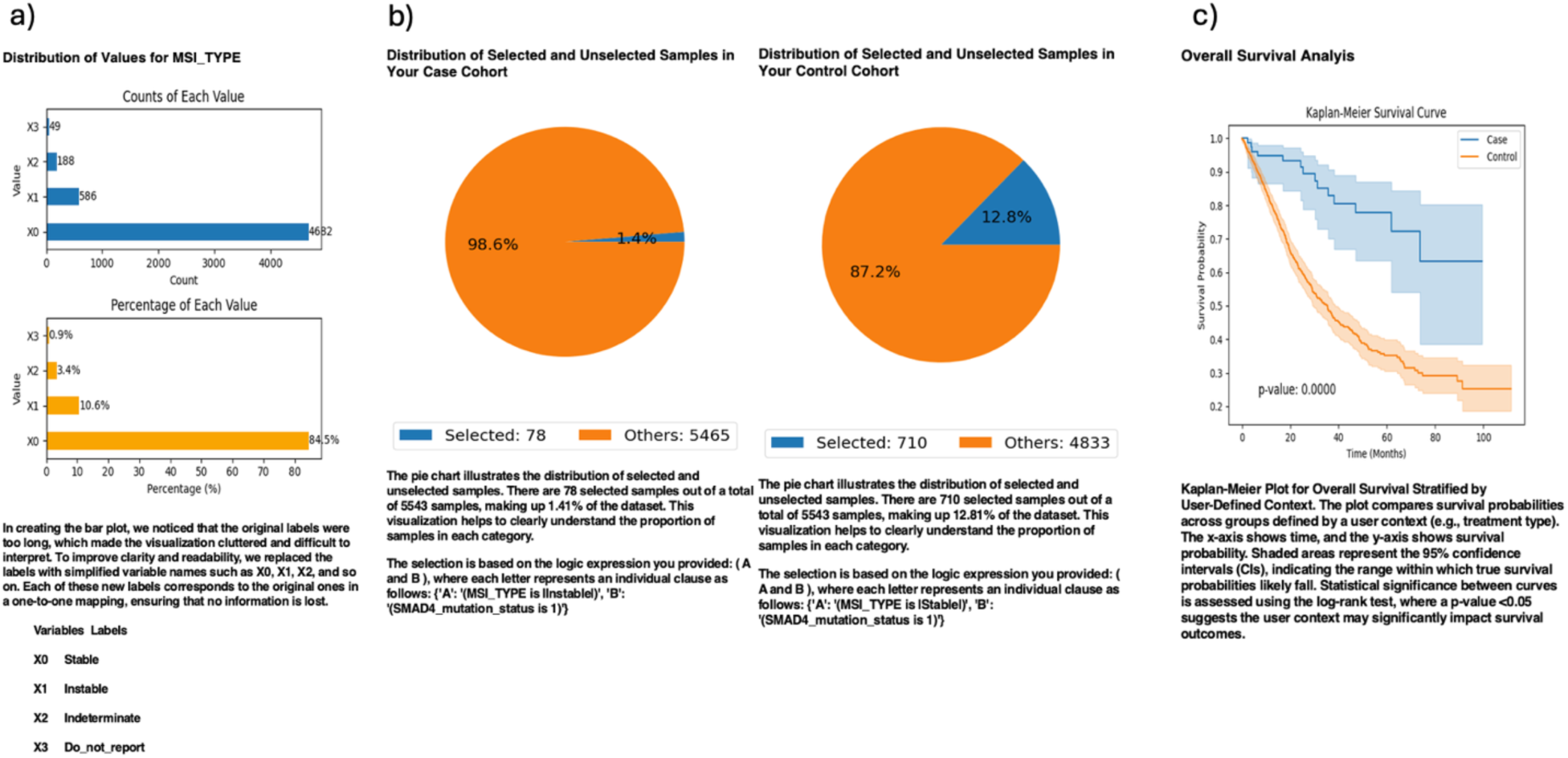
AI-HOPE-TGFbeta Analysis of SMAD4-Mutant Colorectal Cancer (CRC) Patients by Microsatellite Instability Status (MSI-High vs. MSI-Stable). This figure illustrates the application of AI-HOPE-TGFbeta to evaluate survival outcomes in CRC patients with SMAD4 mutations, stratified by microsatellite instability (MSI) status. The case cohort includes patients with MSI-high (Instable) tumors, while the control cohort includes patients with MSI-stable tumors. a) Bar plots summarize the dataset-wide distribution of MSI subtypes. MSI-stable tumors (X0) are the most common, accounting for 84.5% of samples (n=4,662), followed by MSI-instability (X1, 10.6%, n=586). The top chart shows absolute sample counts per MSI category, while the bottom panel displays their proportional representation. Labels were simplified for clarity, with original MSI values mapped to codes (e.g., X0 = Stable, X1 = Instable). b) Pie charts highlight the cohort selection process. The case group (SMAD4-mutated, MSI-Instable) includes 78 samples (1.4% of the total dataset), and the control group (SMAD4-mutated, MSI-Stable) includes 710 samples (12.8%). These charts provide visual context for the relative rarity of MSI-high tumors in the SMAD4-mutant CRC population. c) Kaplan-Meier survival curves compare overall survival between MSI-Instable and MSI-Stable groups within the SMAD4-mutated cohort. Patients with MSI-Instable tumors exhibit significantly improved survival outcomes relative to those with MSI-Stable tumors (p = 0.0000), with a clear divergence between curves and non-overlapping 95% confidence intervals.

**Figure S3.**
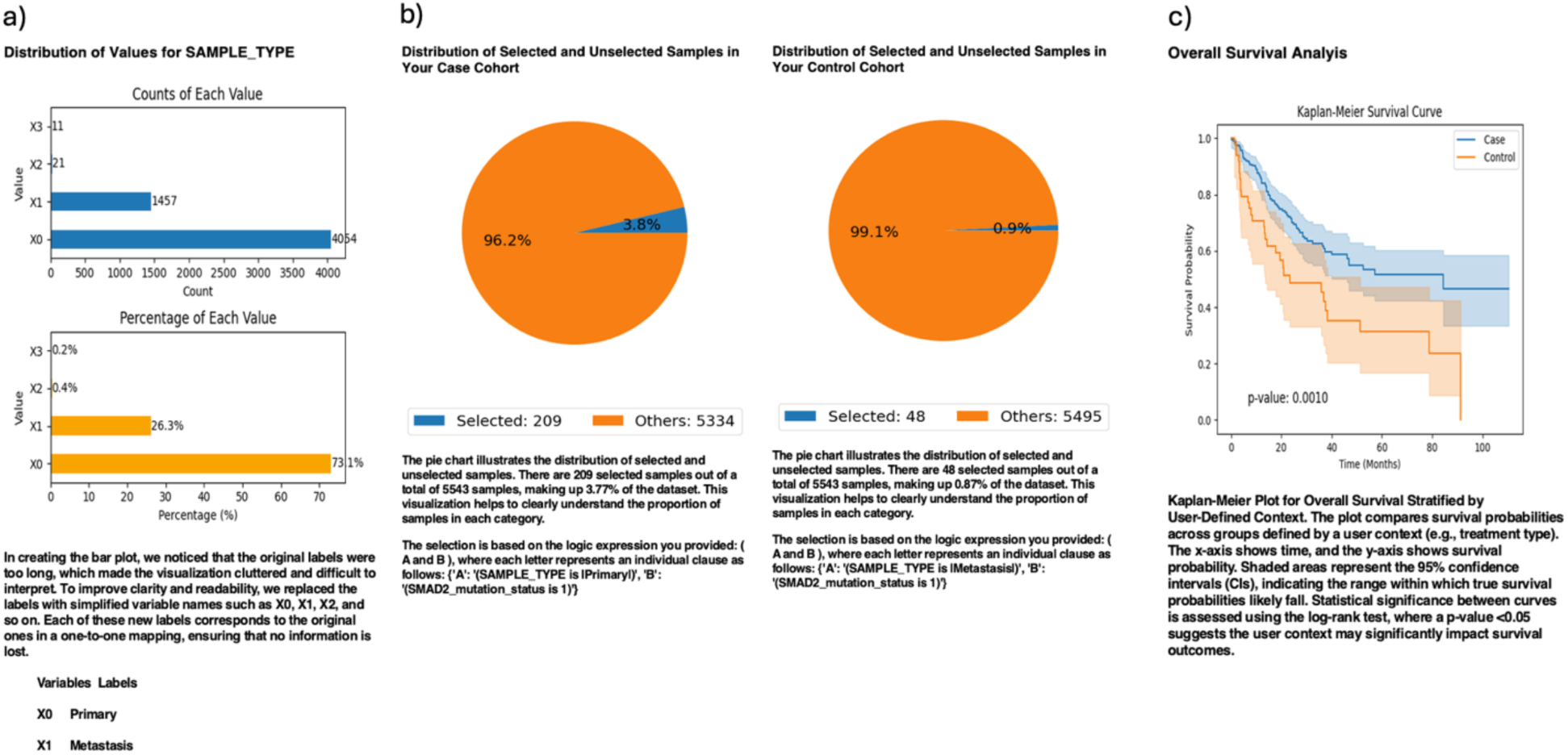
AI-HOPE-TGFbeta Analysis of SMAD2-Mutant Colorectal Cancer (CRC) Patients by Sample Type: Primary vs. Metastatic Tumors. This figure illustrates how AI-HOPE-TGFbeta enables survival analysis of CRC patients with SMAD2 mutations, stratified by tumor sample type (primary vs. metastatic) to assess clinical outcomes across disease progression stages. a) The initial data exploration phase presents bar plots summarizing the distribution of tumor sample types. Primary tumors (X0) represent the majority of samples (n=4,054; 73.1%), while metastatic tumors (X1) account for 26.3% (n=1,457). The upper chart displays absolute counts per group, while the lower chart shows their relative proportions across the dataset. b) Based on the user-defined query, pie charts depict the number of selected samples. The case cohort (primary tumor with SMAD2 mutation) includes 209 samples (3.8% of the dataset), while the control cohort (metastatic tumor with SMAD2 mutation) includes 48 samples (0.9%). These visualizations emphasize the relative prevalence of SMAD2 mutations in primary versus metastatic CRC. c) Kaplan-Meier survival curves compare overall survival between the case and control groups. Patients with SMAD2-mutant primary tumors exhibit significantly better survival than those with SMAD2-mutant metastatic tumors (p = 0.0010), as indicated by distinct curve separation and non-overlapping confidence intervals.

## Notes

### Competing Interest Statement

The authors have declared no competing interest.

### Funding Statement

This work was supported by the National Cancer Institute NCI award number U2CCA252971

### Author Declarations

The source data used in this study were publicly available before the initiation of the study and can be accessed through cBioPortal for Cancer Genomics at https://www.cbioportal.org/ and the GENIE Project (AACR Project GENIE cBioPortal) at https://genie.cbioportal.org. Additional data may be provided upon reasonable request to the authors.

